# Inhaled CO_2_ concentration while wearing face masks: a pilot study using capnography

**DOI:** 10.1101/2022.05.10.22274813

**Authors:** Cecilia Acuti Martellucci, Maria Elena Flacco, Mosè Martellucci, Francesco Saverio Violante, Lamberto Manzoli

## Abstract

None of the available evaluations of the inhaled air carbon dioxide (CO_2_) concentration, while wearing face masks, used professional, real-time capnography with water-removal tubing. We measured the end-tidal CO_2_ using professional side-stream capnography, with water-removing tubing (Rad-97™ capnograph), at rest, (1) without masks, (2) wearing a surgical mask, and (3) wearing a FFP2 respirator, in 102 healthy volunteers aged 10-90 years, from the general population of Ferrara province, Italy. The inhaled air CO_2_ concentration was then computed as: ((mask volume × end-tidal CO_2_) + ((tidal volume - mask volume) × ambient air CO_2_)) / tidal volume).

The mean CO_2_ concentration was 4965±1047 ppm with surgical masks, and 9396±2254 ppm with FFP2 respirators. The proportion of the sample showing a CO_2_ concentration higher than the 5000 ppm acceptable exposure threshold recommended for workers was 40.2% while wearing surgical masks, 99.0% while wearing FFP2 respirators. The mean blood oxygen saturation remained >96%, and the mean end-tidal CO_2_ <33 mmHg. Adjusting for age, gender, BMI, and smoking, the inhaled air CO_2_ concentration significantly increased with increasing respiratory rate (with a mean of 10,143±2782 ppm among the participants taking 18 or more breaths per minute, while wearing FFP2 respirators), and was higher among the minors, who showed a mean CO_2_ concentration of 12,847±2898 ppm, while wearing FFP2 respirators. If these results will be confirmed, the current guidelines on mask-wearing could be updated to integrate recommendations for slow breathing and a more targeted use when contagion risk is low.

## INTRODUCTION

Most nations introduced the use of face masks in the community to contrast SARS-CoV-2 pandemic.(1, 2) Italy, one of the first countries to experience an epidemic wave,(3) required masks to be worn by all people older than five years, indoors and outdoors.(4)

While masks contribute to reducing the epidemic spread,(2) and might also decrease the incidence of other airborne infections,(5, 6) a prolonged mask use has been associated with adverse effects due to pathogens contamination and/or a substantial rise of the carbon dioxide (CO_2_) concentration in inhaled air.(7, 8) However, only two studies directly assessed CO_2_ in air inhaled while wearing masks in the general population.(9, 10) These studies had an overall sample of 12 adults, and used measurement tools which could not avoid the interference of water vapor,(9, 10) which is typically high in exhaled air,(11) and is known to substantially decrease accuracy.(12) Indeed, a third research was recently retracted because of methodological concerns including water vapor interference, instrument’s latency, and the impossibility to correctly define inhaled and exhaled air CO_2_ without the use of capnography.(13)

We used a professional real-time capnograph, with water-removal tubing, in order to assess the inhaled air CO_2_ concentration in a sample of healthy individuals wearing different types of masks.

## MATERIALS AND METHODS

### Study population

The participants were healthy volunteers sequentially recruited by three general practitioners and one family pediatrician in the Province of Ferrara, Italy during April and May 2021. Inclusion criteria were: age between 10 and 90 years, forehead temperature <37.5°C, being able to wear a mask without assistance, and providing written informed consent (for the minors, the consent was requested to the legally responsible individual). Exclusion criteria were: pregnancy, and cardiac or respiratory comorbidities.

### Study design

In this observational, descriptive study, we measured the end-tidal CO_2_ (ETCO_2_) in all participants (a) without masks; (b) wearing a surgical mask; (c) wearing a Filtering Face-Piece grade 2 (FFP2) respirator. Given that the masks constitute an added dead space of the airways, with different volumes depending on mask size and face shape,(14) the concentration of CO_2_ within this added dead space can be assessed measuring the ETCO_2_, which indicates how much CO_2_ is exhaled in the final phase of the expiration.(15) The evaluations of ETCO_2_ were performed after ten minutes of rest, with participants seated, silent, and breathing only through the nose. A trained physician (CAM) took measurements at minutes three, four, and five, and the final value used in the analyses was the average of the three measurements.

All masks were identical and were provided by the investigators, who monitored and eventually adjusted the fit.(16) The surgical mask was a 3-layer plane-shaped disposable face mask with ear loops (17.5×9.5 cm, conforming to UNI EN ISO 14683:2019 and AC:2019 regulations). The FFP2 was a 5-layer disposable respirator (15.0×10.0 cm, conforming to EN 149:2001 and A1:2009), equivalent to United States N95.

The measurement tool was a Rad-97™ capnograph with real-time side-stream gas measurement and water-removal tubing (Masimo Corp., Irvine, CA, USA). The sampling point (nasal cannulas) was positioned outside the exhaled air stream – below the lips of each subject – to ensure that the detected ETCO_2_ was that of the volume of air within the masks. Photos of the measurement method may be obtained from the corresponding author. The capnography device measured CO_2_ in mmHg, which was converted to ppm using a standardized conversion formula.(17)

The environmental CO_2_ concentration (in ppm) was measured using an automatic Temtop mod. M2000C^®^ sensor (Elitech Technology Inc., Milpitas, CA, USA). All measurements were made into a room that was constantly and amply ventilated with external air.

For each participant, information was also collected on blood oxygen saturation and respiratory rate (measured at the same time points as the ETCO_2_), age, gender, weight and height, and smoking (current - at least one cigarette per week - or former). Blood oxygen saturation was measured through a LTD800^®^ digital finger pulse oximeter (Dimed Co. Ltd., Cavriglia, AR, Italy).

### Data analysis

The primary outcome was the mean inhaled air CO_2_ concentration when wearing masks. The secondary outcome was the proportion of individuals with inhaled air CO_2_ concentration exceeding 5000 ppm, which is the long-term occupational exposure (eight hours) threshold recommended by both the United States Department of Labor Occupational Safety and Health Administration (OSHA) and the European Agency for Safety and Health at Work (EU-OSHA).(18, 19)

CO_2_ inhaled air concentration was computed as follows: ((mask volume **×** end tidal CO_2_) + ((tidal volume - mask volume) **×** ambient air CO_2_)) / tidal volume.(20) The standard value of 7 ml per kilogram of weight was used for the tidal volume (the volume of air inhaled and exhaled with every respiration cycle).(21, 22) Similarly, masks volumes were the minimum average values reported by the literature: 50 ml for the surgical mask,(23) and 98 ml for the FFP2 respirator.(24)

The differences in the mean CO_2_ concentration with and without masks were evaluated using Wilcoxon matched-pairs signed ranks test.(25) The analyses were repeated separately for children (aged 10 to 18 years), adults (19-64 years), and elderly (65-90 years), assessing potential differences between the groups through Kruskal-Wallis tests. Multiple linear regression was then performed to investigate potential independent predictors of higher CO_2_ content wearing surgical (model 1) or FFP2 (model 2) masks. All covariates were included a priori in the models, the validity of which was assessed as follows: the assumption of constant error variance was checked graphically, plotting Pearson residuals against fitted values, and formally, using the Cook–Weisberg test for heteroskedasticity. High leverage observations were identified by computing Pearson, standardized and studentized residuals, and Cook’s D influence. We found five high-leverage observations in both models: as their exclusion did not substantially alter the results, the final models were based on the whole sample. A two-tailed p-value<0.05 was considered significant for all analyses, which were carried out using Stata 15.1 (Stata Corp., College Station, TX, 2017).

We decided to enroll a sample size of 100 subjects as it would allow 95% confidence interval to remain within ±10% of the sample mean value, assuming an average inhaled air CO_2_ concentration of 2000±1000 ppm wearing surgical masks, and 3000±1000 ppm wearing FFP2 respirators.(10)

## RESULTS

### Sample characteristics

Participation was requested to 104 eligible subjects; 102 provided the consent and were thus included in the study (50% males; mean age 46.7±19.9 years). Ten participants were aged 10-18 years, 20 were aged 65-90 years. The mean Body Mass Index (BMI) was 24.5±4.6, and current or former smokers were 22.6%. The average respiratory rate was 16.5±3.4 breaths per minute, with 33.3% breathing at or above 18 breaths per minute; the average blood oxygen saturation was 97.4±0.9%, with 98% of the sample at or above 96.0% saturation.

### Outcomes

The mean inhaled air CO_2_ without masks was 458±21 ppm. While wearing the surgical mask, the mean CO_2_ was 4965±1,047 ppm (95% confidence interval 4758 to 5171 ppm), and exceeded 5000 ppm in 40.2% (30.6% to 50.4%) of the measurements (Table 1). While wearing the FFP2 respirator, the average CO_2_ was 9396±2254 ppm (8953 to 9839 ppm), and 99.0% (94.7% to 100%) of the participants showed values higher than 5000 ppm. Among the minors, the mean CO_2_ concentration when wearing surgical masks was 6439±1366 ppm (5462 to 7415 ppm), and was considerably higher than among the adults (4852±857 ppm; p<0.001), or the elderly (4638±948 ppm; p<0.01). A similar difference by age class was observed also for the FFP2 respirators (Table 2). The CO_2_ concentration varied also by respiratory rate: wearing surgical masks, inhaled air CO_2_ was 4663±692 ppm among the individuals with respiratory rate ≤14 breaths per minute, progressively rising to 5271±1291 ppm when 18 or more breaths per minute were taken. A similar trend was observed for FFP2 respirators (Table 1).

**Table 1.** Outcomes for the overall sample and results of the multiple linear regression predicting overall inhaled air CO_2_ in ppm (N = 102).

|  | Without mask | Surgical mask | FFP2 respirator |
| --- | --- | --- | --- |
| *Mean CO<sub>2</sub> detected inside the mask in ppm*<sup>A</sup> |  |  |  |
| Mean±SD (95% CI) | 0±0 (--) | 43 099±4284 (42 257 - 43 940) * | 43 434±4426 (42 565 - 44 303) * |
| *Estimated inhaled air CO<sub>2</sub> in ppm* |  |  |  |
| Mean ± SD (95% CI) | 458±21 (454 - 462)<sup>B</sup> | 4965±1047 (4758 - 5171) * | 9396±2254 (8953 - 9839) * |
| >5000 ppm, % | 0.0 | 40.2 | 99.0 |
| *Inhaled air CO<sub>2</sub> in ppm by respiratory rate, mean±SD (95% CI)* |  |  |  |
| - Slow (≤14 breaths per minute, n=25) | 462±22 (453 - 471) | 4663±692 (4378 - 4950) | 8779±1471 (8171 - 9386) |
| - Moderate (15-17 breaths per minute, n=43) | 457±19 (451 - 463) | 4899±959 (4604 - 5194) | 9165±2043 (8536 - 9793) |
| - High (≥18 breaths per minute, n=34) | 458±22 (450 - 465) | 5271±1291 (4820 - 5721) | 10 143±2782 (9173 - 11 114) |
| *Coefficients for the linear regression* |  |  |  |
| Respiratory rate, 1 breath per minute increase | -- | 33 (-10; 77) | 99 (6; 192)<sup>†</sup> |
| - Low (≤14 breaths per minute, n=25) | -- | 0.00 (Ref. cat.) | 0.00 (Ref. cat.) |
| - Moderate (15-17 breaths per minute, n=43) | -- | 261 (-116; 638) | 431 (-368; 1231) |
| - High (≥18 breaths per minute, n=34) | -- | 546 (157; 935)<sup>‡</sup> | 1243 (418; 2067)<sup>‡</sup> |

FFP2 = filtering face-piece grade 2 respirator. <sup>A</sup>End-tidal CO<sub>2</sub> detected inside the face masks. <sup>B</sup> Only ambient air CO<sub>2</sub>. \* P<0.001 (Wilcoxon matched pairs signed-rank test) of the comparison of CO<sub>2</sub> parameters between without and with surgical or FFP2 masks. <sup>†</sup> P<0.05 and <sup>‡</sup> P<0.01 from the Wald test for the linear regression adjusted by gender, age, Body Mass Index, and smoking status.

**Table 2.**
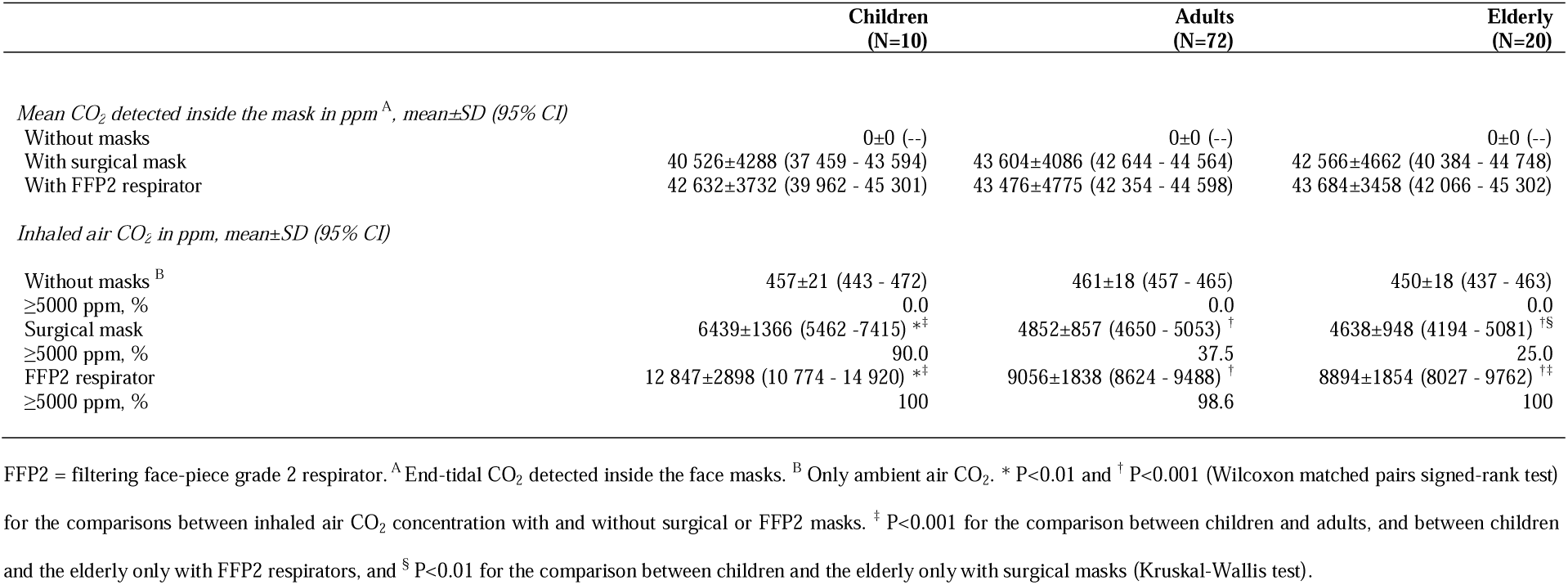
Sample characteristics and outcomes by age-class.

Multivariate analyses substantially confirmed univariate results: a higher respiratory rate was significantly associated with higher inhaled air CO_2_ wearing both masks. Regression coefficients for ≥18 compared to ≤14 breaths per minute were +546 and +1243, respectively, with surgical masks and FFP2 respirators (both p<0.01; Table 1).

Both respiratory rate and blood oxygen saturation did not differ substantially without or with the masks. Also, when wearing masks, the mean ETCO_2_ remained within 33 mmHg.

## DISCUSSION

In our sample of healthy individuals, at rest, after a few minutes of surgical masks use, the mean inhaled air CO_2_ approached the occupational exposure limit of 5000 ppm,(18) and this threshold was largely exceeded when wearing FFP2 respirators. Notably, the CO_2_ concentration significantly increased with increasing respiratory rates, reaching around 5200 ppm in those breathing at 18 or more breaths per minute with surgical masks, and the minors showed substantially higher CO_2_ concentrations than adults.

### Strengths and weaknesses of the study

The chosen capnography device had water-removal tubing, and real-time monitoring, ensuring reliable and reproducible CO_2_ measurements.(26) Indeed, relative humidity ranges 42-91% in exhaled air,(11) potentially altering CO_2_ assessments,(12) which might explain the differences with the measurements of previous studies.(9, 10) Additionally, we examined the largest sample, so far, of healthy individuals of various ages, comparing both surgical masks and FFP2 respirators.(9, 10)

This study has also limitations that must be considered. First, although the sample size is the largest as yet, it is still relatively scarce, especially for the minors. Secondly, the volume of the dead space within the mask could not be assessed for each participant, and therefore we could not closely inspect the possible influence of face shape and individual added dead space on the inhaled air CO_2_. Thirdly, the instrument’s precision of 1.5 mmHg (1974 ppm) widens the uncertainty around the measurements. Importantly, however, when 1974 ppm are subtracted to the mean inhaled air CO_2_, the CO_2_ in surgical masks decreases to about 3000 ppm, while it still exceeds the 5000 ppm threshold with FFP2 respirators.(18) Lastly, the experimental conditions, with participants at complete rest and in a constantly ventilated room, were far from those experienced by workers and students during a typical day, normally spent in rooms shared with other people or doing some degree of physical activity. Since it was observed that speech and even low level physical activity are associated with increases in CO_2_ concentration, CO_2_ values in real life are likely to be higher than those recorded in this study.(27, 28)

### Comparison with other studies and discussion of main findings

High CO_2_ concentrations in masks worn by individuals at rest were previously reported by two studies,(9, 10) which however had very small sample sizes and used instruments that could not avoid the interference of water vapor.(11) The explanation of the observed high CO_2_ values lies in the combination of tidal and mask volumes: even though the 500 ml tidal volume of the average adult man is predominantly filled with low environmental concentrations of CO_2_,(15) the portion represented by the mask dead space had a CO_2_ content so high that the overall inhaled air CO_2_ increased substantially.(29)

Concerning the risk of hypoxia, a research on 53 surgeons found that blood oxygen saturation decreased noticeably with a longer time wearing surgical masks.(29) In contrast, the present study was performed at rest and for a short time, during which the recorded levels of CO_2_ did not substantially alter blood oxygen saturation, as in similar studies.(9, 10, 30) Nevertheless, the exposure to inhaled air CO_2_ values higher than 5000 ppm, for long periods, is considered unacceptable for the workers, and is forbidden in several countries,(18) because it frequently causes signs and symptoms such as headache, nausea, drowsiness, rhinitis, and reduced cognitive performance.(31, 32) Also, reports have been published about the negative impact of respirators on healthcare professionals, such as headache, reduced tolerance to light workload, and recommendations to take regular breaks from mask wearing have been proposed to ensure the wellbeing and productivity of the workers.(33, 34) As regards the difference between mask types, of the above mentioned studies, one did not find differences in the CO_2_ concentration between surgical and FFP2 (with valve) masks, but only subject was analyzed.(10) The other study did not include surgical masks in the evaluation.(9) In fact, given a similar ETCO_2_ between the two mask types, the larger dead space inside FFP2 respirators is expected to determine a sharp difference in CO_2_ content between surgical and FFP2 masks.(23, 24) This is consistent with three previous studies: one on patients whose ETCO2 increased with increasing mask dead space,(35) and two on healthcare professionals (one of which using capnography) which found CO_2_ retention within FFP2 masks, whether with or without valve.(28, 36)

In relation to the respiratory rate, no previous study specifically evaluated its association with CO_2_ concentration in healthy individuals at rest. However, an increase of inhaled air CO_2_ was found during physical activity with masks,(25) and with higher respiratory rates in post-operatory ventilated patients.(37) In addition, it is well known that, besides mask use, slow breathing is associated with significantly lower inhaled air CO_2_ concentration.(38)

Finally, concerning the minors, no study so far directly compared them to adults, and only one relatively old research showed increased CO_2_ concentrations in young children wearing gas masks.(39) In fact, minors can be expected to be at a disadvantage also in this evaluation. Specifically, their small build corresponds to a small tidal volume, which therefore provides lesser dilution of the excess CO_2_ compared to the greater tidal volume of adults.(21, 22) Nonetheless, given the limited number of the included minors, this finding inevitably requires validation.

### Implications of the study

The mentioned OSHA and EU-OSHA 5000 ppm threshold is the long-term Permissible Exposure Limit (eight hours weighted average), while the Short Term Exposure Limit (15 minutes weighted average) is 40 000 ppm.(18, 19) This suggests that limiting mask use to short intervals, for instance when visiting shops, does not imply an immediate risk of inhaling excess CO_2_. Instead, if the CO_2_ concentration measured in the present work is confirmed during more protracted mask-wearing, the use of masks could be targeted to the sites and hours in which SARS-CoV-2 transmission risk is moderate to high, and reduced as much as possible when the risk is low, especially in view of the rising SARS-CoV-2 vaccination coverage worldwide.(40) Indeed, given the recent evidence suggesting that, in crowded rooms, two air-changes per hour may lower aerosol build-up more efficiently than the best performing masks,(41) the choice of increased ventilation over mask-wearing could be taken into consideration when allowed by the environmental and epidemiological conditions, especially for the minors.

Moreover, the observed difference in inhaled air CO_2_ according to mask types suggests that, when the usage is protracted and/or physical activities are required, surgical masks should be used instead of FFP2 respirators, as they reduce the possible negative effects of high CO_2_ concentrations, while still covering nose and mouth and therefore reducing the emission of droplets and aerosols.(42, 43) Finally, if the relationship between CO_2_ levels and respiratory rate is verified, the current guidelines to control SARS-CoV-2 pandemic could be integrated with recommendations to reduce the respiratory rate during mask use.(38) This would be particularly important for blue-collar workers and the elderly, who are known to breathe faster,(44, 45) possibly more so when wearing masks,(7) and showed higher baseline CO_2_ concentrations.(46) Indeed, the average respiratory rate at rest has been estimated around 15 breaths per minute in healthy adults,(47) and, in the present assessment, three additional breaths per minute were enough to increase the mean CO_2_ content over 5000 ppm when wearing surgical masks.

### Unanswered questions and future research

Mask wearing is required in many countries throughout the working day, and during lectures in the case of students.(2) Therefore, capnography and pulse oxymeter monitoring of a general population sample should be extended to more than five minutes, possibly to hours of observation, and not only in conditions of rest, in order to verify whether the ETCO_2_ detected within masks remains constant or has a tendency to increase with longer mask wearing and while performing habitual tasks. In addition, subjective symptoms such as headache and drowsiness should also be investigated.

As mentioned, the progressive rise in CO_2_ with increasing breaths per minute, and the higher CO_2_ in minors also requires validation from further studies with larger samples.

## CONCLUSIONS

Shortly after wearing surgical masks, the inhaled air CO_2_ approached the highest acceptable exposure threshold recommended for workers, while concerningly high concentrations were recorded in virtually all individuals when wearing FFP2 masks. The CO_2_ concentration was significantly higher among minors and the subjects with high respiratory rate. If these findings are confirmed, the current guidelines on masks use could be updated to integrate recommendations for slow breathing and a more targeted use when contagion risk is low.

## Data Availability

The study data is available from the corresponding author upon reasonable request.

## Acknowledgements

The authors thank Luisa Rogari and Francesca and Marta Rosini for their help in data collection.

## Ethical approval

The study protocol was approved by the Ethics Committee of the Emilia-Romagna Region “Area Vasta Emilia Centrale” on February 12^th^ 2021 (code 78/2021/Oss/UniFe).

## Data availability

The study data is available from the corresponding author upon reasonable request.

## FOOTNOTES

### Contributors

CAM and LM: conceptualization and methodology. CAM, MEF, and MM: investigation. LM: funding acquisition and project administration. CAM and MEF: formal analysis. CAM, MEF, MM, FSV, and LM: data curation. LM and FSV: supervision and validation. CAM, MEF, and LM: writing - original draft. CAM, LM, and FSV: writing - review and editing. All authors critically revised the article for important intellectual content and gave final approval for the article. The corresponding author attests that all listed authors meet authorship criteria and that no others meeting the criteria have been omitted.

